# Exploring the global impact of the COVID-19 pandemic on medical education: an international cross-sectional study of medical learners

**DOI:** 10.1101/2020.09.01.20186304

**Authors:** Allison Brown, Aliya Kassam, Mike Paget, Kenneth Blades, Megan Mercia, Rahim Kachra

## Abstract

To broadly explore the extent that COVID-19 has initially impacted medical learners around the world and examine global trends and patterns across geographic regions and levels of training, a cross-sectional survey of medical learners was conducted between March 25-June 14^th^, 2020, shortly after the World Health Organization declared concurrent COVID-19 a pandemic. 6492 medical learners completed the survey from 140 countries, Students were concerned about the quality of their learning, training progression, and milestone fulfillment. Most trainees felt under-utilized and wanted to be engaged clinically in meaningful ways; however, some trainees felt that contributing to healthcare during a pandemic was beyond the scope of a medical learner. Statistically significant differences were detected between levels of training and geographic regions for satisfaction with organizational responses, the impact of COVID-19 on wellness, and state-trait anxiety. Overall, the initial disruption to medical training has been perceived by learners across all levels and geographic regions to have negatively affected their training and well-being, particularly amongst postgraduate trainees. These results provide initial insights into the areas that warrant future research as well as consideration for current and future policy planning, such as the policies for clinical utilization of medical learners during public health emergencies.

## Introduction

The coronavirus pandemic (COVID-19) represents the first global interruption to medical education since World War II. This public health emergency has required undergraduate and postgraduate medical training programs around the world to rapidly respond, including re-deploying learners to other clinical spaces or removing them entirely, and moving education online to promote physical distancing (Association of American Medical Colleges 2020; Harvey 2020; Rose 2020). The current COVID-19 pandemic has changed the status quo of medical training as we know it – whether for better or for worse remains unclear and may have profound implications.

Prior localized disruptions to medical training have negatively affected trainees, such as pandemics and climate emergencies. The severe acute respiratory syndrome (SARS) pandemic in 2003 revealed the tension between embracing learning opportunities that a public health emergency offers and protecting the needs and well-being of learners (Rieder et al. 2004; Sherbino & Atzema 2004; Rambaldini et al. 2005). The SARS pandemic adversely affected the mental health of learners and healthcare professionals, and heightened anxiety amongst learners was associated with a perceived negative educational experience (Maunder et al. 2003; Bai et al. 2004; Landis & Bradley 2005; Rambaldini et al. 2005). In 2005, the rapid restructuring of the medical school curriculum in response to Hurricane Katrina resulted in a decline in academic performance (Crawford et al. 2008). Lessons from both SARS and Hurricane Katrina highlighted the importance of support networks and attention to learner wellness, recognizing the additional stressors that these disruptions can place on medical trainees (Landis & Bradley 2005; Rambaldini et al. 2005; DiCarlo et al. 2007; Crawford et al. 2008). These emergencies led to significant and rapid innovation in curriculum structure and content (DiCarlo et al. 2007). While disruptions to medical education can provide an impetus to reconsider ethical and practical implications of training during emergencies, training models may remain stagnant (Landis & Bradley 2005; DiCarlo et al. 2007). Despite a heightened awareness fifteen years ago of the potential implications of interruptions to medical education, many organizations and systems entered the current pandemic relatively unprepared.

Medical education’s understanding of the impact of global emergencies and disruptions to medical training remains limited to SARS and localized emergencies, yet the current pandemic remains the single most substantial disruption to contemporary medical training with global impact. The extent to which COVID-19 has impacted medical training remains speculative and anecdotal, and the consequences and implications may be profound. The research question for this exploratory study was: How have medical learners been impacted by the COVID-19 pandemic around the world?

## Methods

### Data Collection

We administered a cross-sectional survey between March 25 and June 14, 2020, shortly after the World Health Organization declared COVID-19 a pandemic. Criterion and snowball sampling techniques were used to collect data from undergraduate (e.g., medical students) and postgraduate (e.g., interns, house staff, resident physicians) medical learners at any medical school around the world. This study explicitly focused on medical learners given that they provide a significant proportion of patient care yet are still considered learners.

Our decision to use a survey was based on the ability of this method to efficiently collect empirical data from a large population (Kelley et al. 2003) The survey instrument was designed to include both quantitative and qualitative questions in order to later compare variables across groups, allow participants to provide insight into how they have been impacted using their own words, and use the qualitative data to potentially expand upon the results of the quantitative data (Appendix A). All participants completed demographic questions followed by questions specific to their level of training. Next, shared questions explored the effect of the pandemic on learner wellbeing and their communications with others. Open-ended questions collected qualitative data surrounding the impacts of the pandemic on training, utilization of residents, and strategies used in response to the pandemic. All questions were developed *de novo*, except the 6-item State-Trait Anxiety Inventory (STAI-6) (Marteau & Bekker 1992). One question exploring the impact of the pandemic on five domains of wellness was informed by a holistic framework for learner wellness (Kassam & Ellaway 2020).

An initial survey underwent pilot testing to refine comprehension and functionality. The final survey was translated to 19 additional languages to increase the inclusion of learners from around the world. The survey was administered online using Qualtrics software by distributing an anonymous, re-usable link over social media and through snowball sampling.

### Data Analysis

Quantitative data were imported into SPSS Version 26 (IBM Corp., New York) for analysis. Countries were coded to the geographic regions defined by the World Bank to explore geographic differences (World Bank) Descriptive statistics were calculated for each item. Total scores were computed for scaled questions, with positively worded items reverse coded in the calculation. Independent t-tests were used to compare total scores between medical students and residents, and an Analysis of Variance test was used to compare data across geographic regions using a post-hoc correction to account for multiple comparisons. To examine the psychometric properties of the survey, Cronbach’s alpha was computed for scaled items to assess reliability evidence and an exploratory factor analysis was used to examine construct validity.

Qualitative data were back translated to English and analyzed in NVivo Version 12 (QSR International, Melbourne). Three investigators read through the data to identify emerging themes, which were then organized into a coding framework for deductive thematic and content analysis to similarly examine trends between levels and across geographic regions (Miles et al. 2014). Two investigators (KB, MM) independently coded 100 responses to refine the initial coding framework and assess inter-rater agreement. After an additional 400 responses were analyzed and the Kappa statistic continued to suggest acceptable inter-rater agreement, and a decision was made to divide to improve efficiency given the volume of data. Upon completion of the coding for all responses, themes in the coding framework were divided amongst the research team. Each member analyzed the data coded to their assigned themes and summarized the trends and patterns in the data. Theme summaries were compiled for interpretation by the study team, who then compared and contrasted the results of the qualitative data with the quantitative data to examine how these findings converge or diverge from, and ultimately expand upon, the quantitative results.

The University of Calgary Conjoint Health Research Ethics Board reviewed and approved this study (File #20-0484). Participation in this study was voluntary. Implied consent was obtained from all participants. Participants had the option to either skip individual questions or select “prefer not to answer.”

## Results

A total of 6492 learners from 140 countries completed the survey. The majority of participants were medical students (81.0%) and from Europe and Central Asia (43.4%) (Table 1). The results for both medical student-specific and resident-specific questions are summarized in Table 2. Figure 1 visualizes the responses to the scaled questions for both levels (see Appendix B for additional visualizations of the data by level of training and geographic region).

**Table 1.**
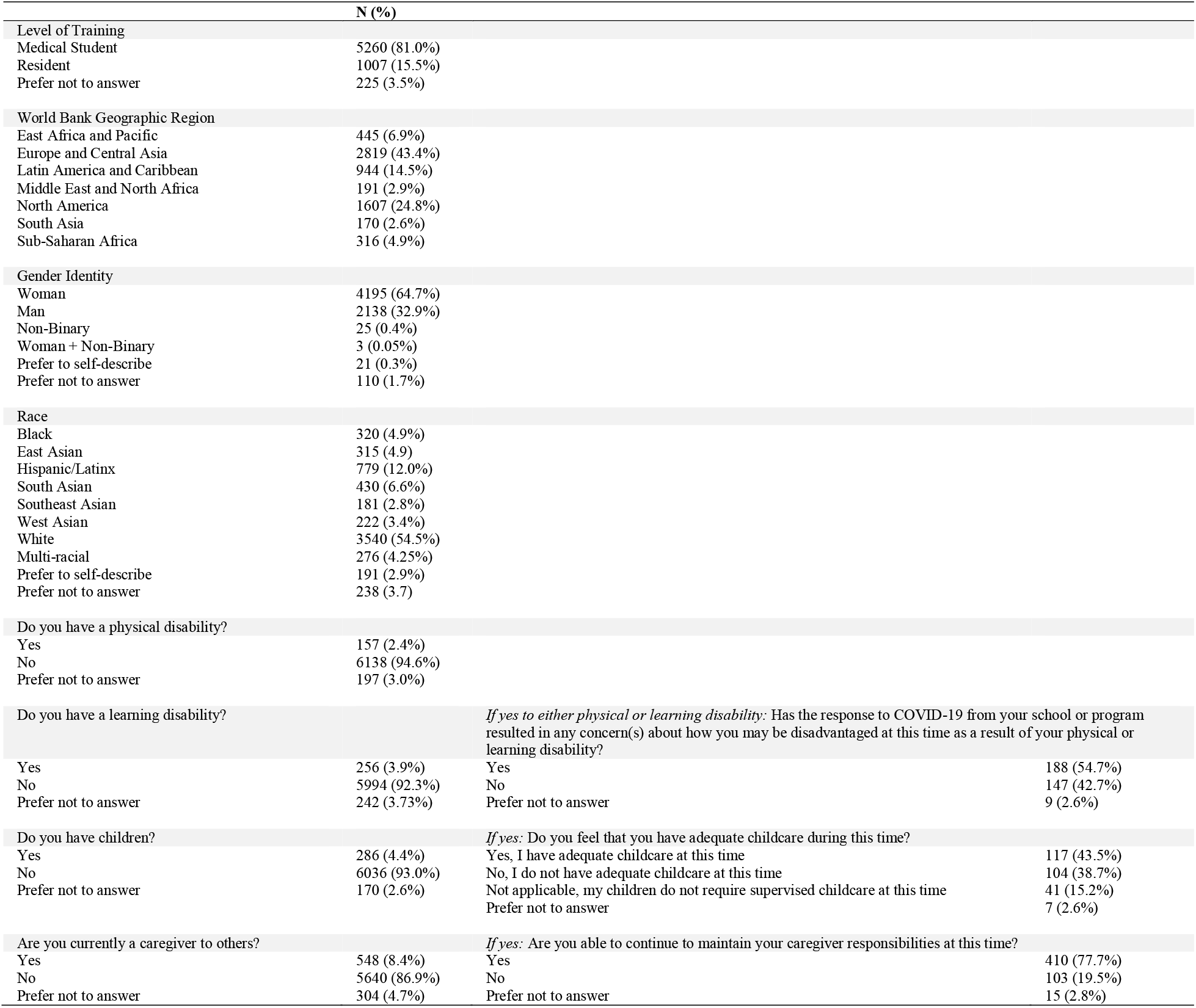
Demographic Characteristics of Participants

**Table 2.** Medical Student and Resident Specific Questions

| Medical Students (N=5260) | N (%) |
| --- | --- |
| *Clinical level students only:* Did your medical school formally excuse you from clinical duties (i.e., clerkship rotations, other clinical rotations or experiences) as a result of the COVID-19 pandemic?* (N=2426 clinical level students) |  |
| Yes | 2044 (84.3%) |
| No | 231 (9.5%) |
| Prefer not to answer | 151 (6.22%) |
| Are you currently working clinically? |  |
| Yes, I am currently working in my planned clinical rotations as a medical student | 246 (4.7%) |
| Yes, I am currently working clinically but in other ways at this time | 227 (4.3%) |
| No, I am not working clinically | 2033 (38.7%) |
| No, I am not working clinically because I am in the pre-clinical stage of training | 2483 (47.2%) |
| Prefer not to answer | 271 (5.1%) |
| If your medical school has excused you from clinical duties or moved your scheduled curriculum to online learning modalities, did you leave or depart from your primary residence (i.e., where you live during the school year) to go to another location during this time? ** |  |
| Parent(s) or family member’s house | 2028 (38.6%) |
| Friend’s house | 58 (1.1%) |
| Somewhere else | 132 (2.5%) |
| Did not leave | 2624 (49.9%) |
| Prefer not to answer | 135 (2.6%) |
| How are you spending your time at the moment?*** |  |
| My learning/training has not been disrupted so I am spending my time as I typically would during the medical school year | 757 (14.4%) |
| My learning is now online as coordinated by the medical school | 3587 (68.2%) |
| I am engaging in self-directed learning | 2322 (44.1%) |
| I am studying for licensing examinations | 838 (15.9%) |
| I am working on research or scholarly projects | 887 (16.9%) |
| I am volunteering | 622 (11.8%) |
| I am helping with contact tracing | 140 (2.7%) |
| I am taking this time to relax or rest | 1913 (36.4%) |
| I am taking care of someone who is ill | 123 (2.3%) |
| I am personally unwell and need to self-isolate | 121 (2.3%) |
| I am spending this time taking care of my children/family | 437 (8.3%) |
| I am taking this time to engage in more wellness activities than I usually would | 1654 (31.4%) |
| Other | 267 (5.1%) |
| Prefer not to answer | 26 (0.5%) |
| *If Yes to volunteering:* How are you currently volunteering?* |  |
| Contact tracing | 102 (1.9%) |
| Call centres | 122 (2.3%) |
| Screening centres | 64 (1.2%) |
| Providing child care to healthcare professionals | 79 (1.5%) |
| Providing child care to others | 27 (0.5%) |
| Other | 357 (6.8%) |
| Prefer not to answer | 19 (0.4%) |
| Have you participated in any online learning during this time that was coordinated or organized by your medical school for aspects of your core curriculum (e.g., lectures or clerkship teaching sessions on Zoom)? (N=4938 valid responses) |  |
| Yes | 3997 (80.9%) |
| No | 914 (18.5%) |
| Prefer not to answer | 27 (0.5%) |
| *If yes to Online learning:* Do you feel that the quality of your medical education will be lower as a result of any online learning your school is coordinating at this time? (N=3919 valid responses) |  |
| Yes | 2608 (66.5%) |
| No | 1225 (31.3%) |
| Prefer not to answer | 86 (2.2%) |
| *If yes to Online learning:* Do you have any challenges accessing online learning modalities (e.g., Zoom, online lectures)? (N=3919)** |  |
| I have difficulties accessing a computer | 89 (2.0%) |
| I have difficulties accessing the internet | 431 (11.0%) |
| I have difficulties learning from home | 835 (21.0%) |
| I have other difficulties | 202 (5.0%) |
| No, I do not have any difficulties accessing online learning | 2743 (70.0%) |
| Prefer not to answer | 31 (1.0%) |
| How has the COVID-19 pandemic directly impacted your clinical electives? |  |
| My visiting electives at other institutions have been cancelled or postponed at this time | 1027 (19.5%) |
| My core electives at my own medical school have been cancelled or postponed at this time | 1235 (23.5%) |
| My electives have not yet been influenced at this time | 203 (3.9%) |
| Not applicable, as all of my electives have been completed | 213 (4.0%) |
| Prefer not to answer | 110 (2.1%) |
| How has the COVID-19 pandemic influenced your career decision-making (e.g., residency interests, career planning)?<sup>a</sup> |  |
| It has influenced the residency program(s) I wish to apply to | 564 (10.7%) |
| It has increased my interest in Public Health as a specialty | 600 (11.4%) |
| It has increased my interest in Infectious Diseases as a specialty | 600 (11.4%) |
| It has increased my interest in Family Medicine as a specialty | 314 (6.0%) |
| It has increased my interest in other programs (open-ended) | 268 (5.1%) |
| It has not influenced my career decision making or interests | 2944 (56.0%) |
| Prefer not to answer | 130 (2.5%) |
| Residents (N=1148 respondents for resident-specific items) | N (%) |
| Setting |  |
| Urban | 766 (66.7%) |
| Rural | 121 (10.5%) |
| Other | 23 (2.0%) |
| Prefer not to answer | 238 (20.7%) |
| Program |  |
| Anesthesiology | 50 (4.4%) |
| Emergency Medicine | 30 (2.6%) |
| Family Medicine | 142 (12.3%) |
| Internal Medicine (core) | 147 (12.8%) |
| Medical Subspecialty | 38 (3.3%) |
| Neurology | 11 (1.0%) |
| Obstetrics and Gynecology | 45 (3.9%) |
| Pediatrics | 42 (3.6%) |
| Public Health | 28 (2.4%) |
| Psychiatry | 42 (3.6%) |
| Radiology | 19 (1.6%) |
| Surgery | 113 (9.8%) |
| Other | 290 (25.3%) |
| Prefer not to answer |  |
| Are you working more than usual during the COVID-19 pandemic? |  |
| Yes, I am working more than usual | 184 (16.0%) |
| No, I am working the same amount | 254 (22.1%) |
| No, I am working less than usual | 421 (36.7%) |
| Prefer not to answer | 289 (25.1%) |
| *FM or Psychiatry Residents:* Do you think residents in these programs could potentially be utilized at this time to provide peer-to-peer mental health support and/or counseling to other residents or healthcare professionals at this time over videoconferencing modalities (N=184) |  |
| Yes, I think we could be useful to provide mental health supports at this time to any and all other residents at this time | 89 (48.4%) |
| Yes, I think we could be useful to provide mental health supports at this time to residents who are working at the front lines of care, or who may be additionally stressed or burnt out as a result of the pandemic | 66 (35.9%) |
| Yes, I think we could be useful to provide mental health supports at this time to other healthcare professionals (e.g., nursing) at this time | 50 (27.2%) |
| No, I do not think we could be useful at this time in providing mental health supports to other residents | 51 (27.7%) |

<sup>a</sup>participants could select all options that applied, % = of all respondents at that level

**Figure 1.**
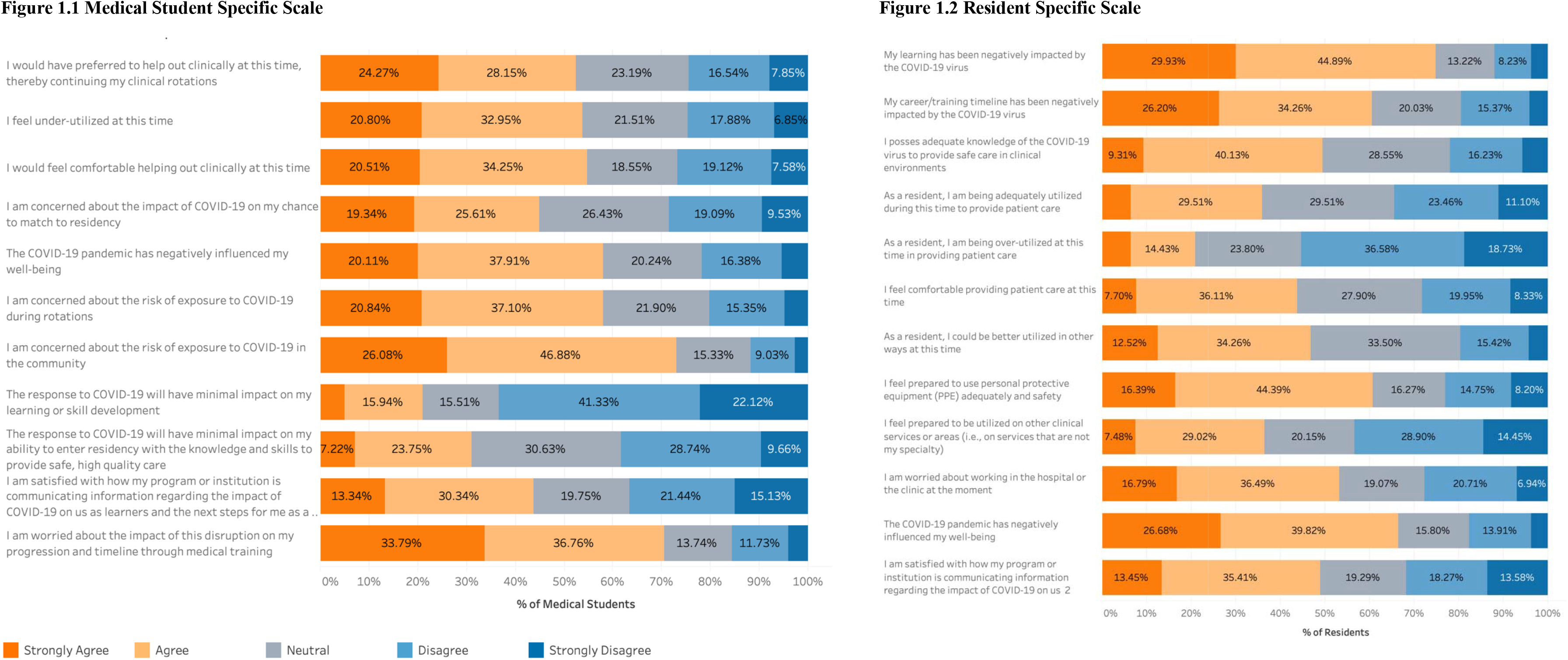
Scaled Questions Exploring the Impact of COVID on Learners

### Undergraduate Medical Learners

The majority (84.3%) of clinical-level medical students were excused from clinical duties; however, some worked clinically in some capacity (9.0%). Most spent their time in online learning coordinated by their medical school (68.2%), pursuing self-directed learning (44.1%), resting (36.4%), and engaging in more wellness activities than normal (31.4%). Most (80.9%) had online learning coordinated by their school but 66.5% felt their education would be of lower quality as a result. Some had difficulties learning from home (21.0%) and accessing the internet (11.0%).

Students reported that their core electives (23.5%) and visiting electives (19.5%) were cancelled or postponed. The qualitative data highlighted how reduced clinical exposure limited opportunities to explore specialties, decreased career interests, and created anxiety about implications for future milestones (e.g., examinations, residency match). While the majority of medical students did not report the pandemic influenced their career interests, 11.4% of medical students reported increased interests in both Public Health and Infectious Disease as a result of the pandemic, as well as other clinical specialties (e.g., Emergency Medicine, Critical Care, Internal Medicine) and new interests in incorporating epidemiology into their future careers. Decreased interest in clinical careers and concerns about the impact of the pandemic on job prospects in their country were noted qualitatively.

Over half of medical students reported feeling under-utilized (53.8%); 52.4% wanted to assist clinically (52.4%) and felt comfortable doing so (54.8%). While 57.9% worried about the risk of exposure to the virus during rotations, 73.0% worried about the risk of exposure in the community. Over two thirds (70.6%) reported feeling worried about the disruption to their progression through medical training, with 45.0% worrying about their chances to match to residency. Only 21.0% thought the pandemic would have a minimal impact on their learning or skill development, and only 31.0% felt that it would have a minimal impact on their ability to enter residency with sufficient knowledge and skill to provide safe, high-quality care. The majority of students (58.0%) felt that the pandemic negatively affected their wellbeing.

### Postgraduate Resident Learners

Residents were from a variety of specialties, mostly in urban settings. Three-quarters of residents agreed their learning was negatively impacted by COVID-19 and 60% felt it disrupted their career progression and timeline. Only 16% reported working more than normal during the pandemic and 36.7% working less than normal. Some residents felt adequately utilized (35.9%) or over-utilized (20.9%); however, 46.8% reported they could be better utilized. Over one-third felt comfortable providing patient care at this time, and 60.8% felt prepared to use personal protective equipment (PPE). Half of residents reported adequate knowledge of COVID-19 to provide care, 36.5% were prepared to be redeployed to other clinical services, and only 53% were worried about working in the hospital. 66.5% agreed that COVID-19 had negatively impacted their wellbeing. The qualitative data provided further evidence that residents felt under-utilized - but highlighted how they saw their re-deployment in a clinical setting as being in conflict with their educational goals.

### Shared Questions

All scaled items had acceptable internal consistency (α>.7) and factor loadings, suggesting reliability and validity evidence. Statistically significant differences were detected between levels of training (p<.05) and geographic regions (p<.007) for the items examining satisfaction with organizational responses [t=7.517, p<.000; [F(6,5258) = 73.85, p<.007]], the impact of COVID-19 on wellness [t =-7.774, p<.000 [F(6,5165)= 11.81, p<.007]], and state-trait anxiety [t=-4.710, p<.000;[F(6, 5131)=10.48, p<.007]] (Appendix C). In general, residents reported lower satisfaction with organizational responses (x=15.67, 95% CI = 15.36-159.98) as well as a greater negative impact on their wellbeing (x=18.52, 95% CI 18.24-18.80,) and anxiety (x=15.13, 95% CI 14.83-15.42). Learners from the East Asia and Pacific region were most satisfied with organizational responses, whereas learners from Sub-Saharan Africa and Latin America and Caribbean were least satisfied (Figure 2.1). Medical students from the Latin America and Caribbean region as well as residents from North America reported the largest negative impact of COVID-19 on their wellness (Figure 2.2). Residents from South Asia, Latin America and Caribbean reported the highest state-trait anxiety (Figure 2.3).

**Figure 2:**
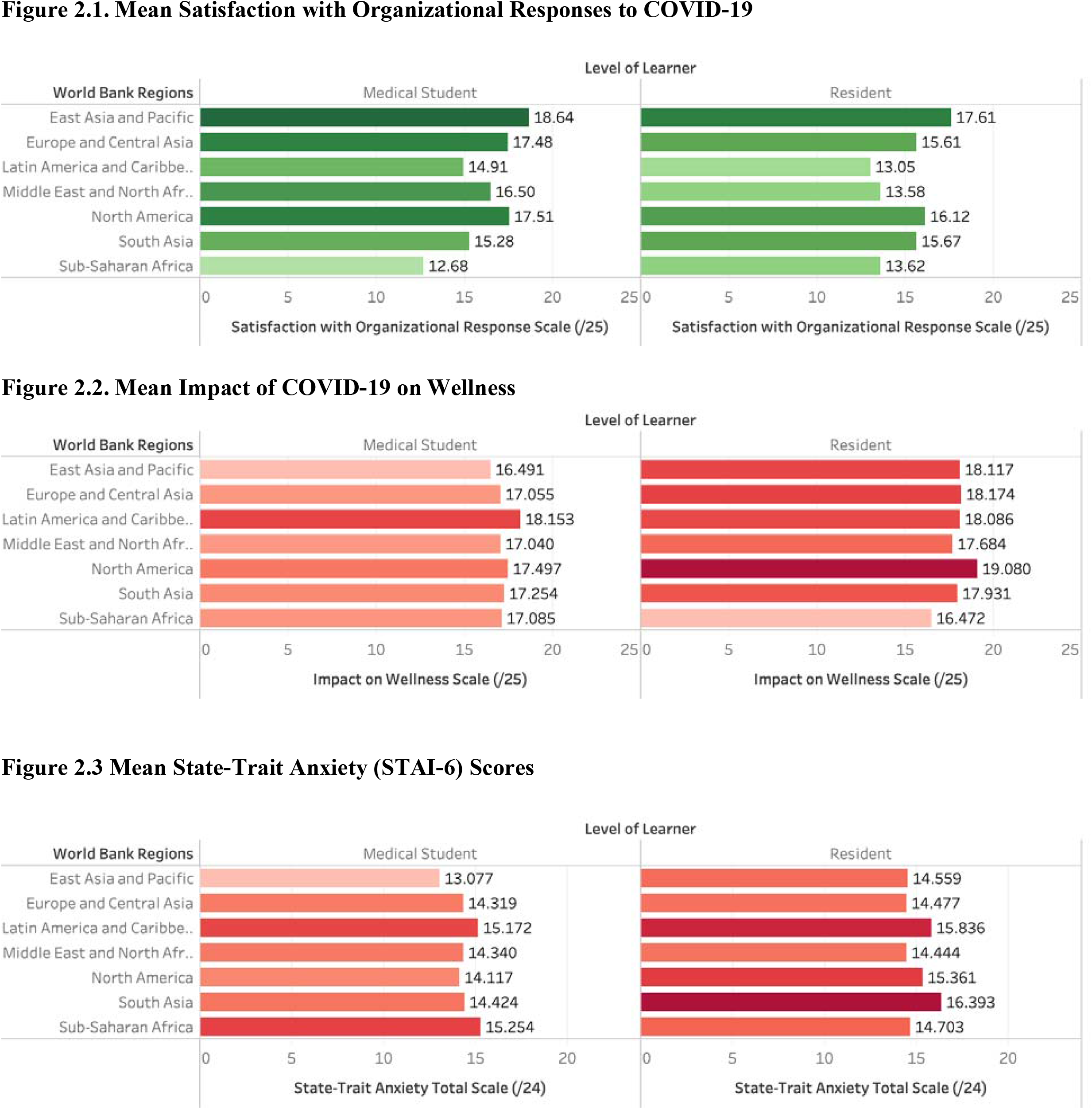
Comparison of Shared Scaled Items Between Levels and Geographic Regions

Seventy-four percent of residents and 57% of medical students reported being more anxious than usual (Table 3; Figure B.4). Common concerns were the health and wellbeing of family members, the impact of the pandemic on their learning, the health of the public, their personal health, the financial situation of others, and their personal financial situation (Figure B.3). One-quarter of participants identifying as Asian reported an increase in racist comments or behaviors in the clinical setting during COVID-19. Over half (54.7%) of learners living with a disability felt their school’s response to the pandemic would further disadvantage them due to their disability.

**Table 3.** Shared Questions

|  |  |  |  |  |
| --- | --- | --- | --- | --- |
| *Participants who selected any Asian race category:* Have you noticed or experienced an increase in racist comments or behaviors towards you or other Asian individuals in the clinical setting since the emergence of COVID-19?<br>(N=944) |  |  |  | Total (N=944) |
| Yes |  |  |  | 231 (24.5%) |
| No |  |  |  | 631 (66.8%) |
| Prefer not to answer |  |  |  | 82 (8.7%) |
| Which of the following are concerns or stressors for you at this time?<sup>b</sup> | Medical Student | Resident | Did not specify level | Total |
| My personal health and well-being | 2281 | 440 | 58 | 2779 |
| The health and well-being of my family members | 3518 | 605 | 87 | 4210 |
| The health and wellbeing of the public | 2761 | 509 | 72 | 3342 |
| My personal financial situation | 1391 | 200 | 51 | 1642 |
| The financial situation of others | 2089 | 339 | 57 | 2485 |
| The impact of this pandemic on my learning | 3046 | 468 | 71 | 3585 |
| Other | 253 | 83 | 9 | 345 |
| Would you consider yourself more anxious during this time than you typically are during the year? | Medical Student (N=4361) | Resident (N=737) | Did not specify level (N=131) | Total (N=5229) |
| Yes | 2505 (57.4%) | 547 (74.2%) | 65 (49.6%) | 3117 (59.6%) |
| No | 1779 (40.8%) | 181 (24.6%) | 51 (38.9%) | 2011 (38.5%) |
| Prefer not to answer | 77 (1.8%) | 9 (1.2%) | 15 (11.5%) | 101 (1.9%) |
| Are you practicing social or physical distancing? | Medical Student (N=4367) | Resident (N=742) | Did not specify level (N=132) | Total (N=5241) |
| Almost always | 3206 (73.4%) | 430 (58.0%) | 72 (54.5%) | 3708 (70.7%) |
| As much as I can | 1099 (25.2%) | 293 (39.5%) | 51 (38.6%) | 1443 (27.5%) |
| Not at all | 49 (1.1%) | 13 (1.8%) | 4 (3.0%) | 66 (1.3%) |
| Prefer not to answer | 13 (0.3%) | 6 (0.8%) | 5 (3.8%) | 24 (0.5%) |
| Have you had to quarantine or self-isolate? | Medical Student (N=4360) | Resident (N=741) | Did not specify level (N=130) | Total (N=5231) |
| Yes | 596 (13.7%) | 160 (21.6%) | 19 (14.6%) | 775 (14.8%) |
| No | 3749 (86.0%) | 574 (77.5%) | 103 (79.2%) | 4426 (84.6%) |
| Prefer not to answer | 15 (0.3%) | 7 (0.9%) | 8 (6.2%) | 30 (0.6%) |
| Are you being asked by others to provide medical expertise or information regarding the COVID-19 pandemic?<sup>b</sup> | Medical Student | Resident | Did not specify level | Total |
| Yes – Family | 2477 | 529 | 71 | 3077 |
| Yes – Friends | 1965 | 452 | 60 | 2477 |
| Yes – Others | 591 | 180 | 23 | 794 |
| No | 1392 | 111 | 38 | 1541 |
| Prefer not to answer | 46 | 10 | 5 | 61 |
| Are you using social media to communicate to others about COVID-19?<sup>a</sup> | Medical Student | Resident | Did not specify level | Total |
| Yes – Twitter | 506 | 123 | 25 | 654 |
| Yes – Instagram | 1065 | 135 | 37 | 1237 |
| Yes – Facebook | 1208 | 247 | 38 | 1493 |
| Yes - Other | 352 | 78 | 25 | 455 |
| No | 2220 | 347 | 45 | 2612 |
| Prefer not to answer | 58 | 9 | 11 | 78 |

<sup>a</sup>N=respondents for each question; questions were not mandatory so total number of responses per question does not equal the total number of participants throughout the study; <sup>b</sup>“select all that apply” question, no proportions calculated

Some learners reported having to be in quarantine or isolation at the time of the survey. Nearly all learners were practicing physical and social distancing.

The qualitative data highlighted how normal education and training processes were severely disrupted by the pandemic (Appendix D). Learners identified a need for institutions to adapt and innovate in order to mitigate these impacts and continue education. Learners also desired to have input into their institutions’ pandemic response. Strategies such as town halls, surveys, and learner representation on decision-making committees helped ensure their circumstances were accurately understood and enabled them to raise concerns overlooked by decision-makers.

Clear and effective communication during the pandemic was essential for learners, while poor or absent communication left learners uncertain of what was happening and contributed to their anxiety. Effective communication provided clarity and reassurance about the pandemic’s impacts and the institutional response. Learners wished to be informed about impacts on their programs, details of the pandemic response, their role (including how to help), and resources they could access. Effective communications about these topics were clear, specific, decisive, concise, and reassuring. Irregular (sporadic) communication created an “information vacuum” that was filled by speculation, rumors, and misinformation. Overly frequent communication was overwhelming and led students to ignore institutional communications.

Numerous interventions were implemented in response to COVID-19, including virtual teaching strategies (e.g., Zoom, Microsoft Teams), office hours and town halls, and clinical strategies (e.g., patient handover via teleconference, alternating call schedules to minimize exposure). When done effectively, online learning maintained educational momentum and provided learners with motivation, a routine, and a sense of purpose. Self-directed learning was frequently used to augment online learning, and when provided with support and guidance, this motivated learners and provided them with a sense of progress. Programs with too much content overwhelmed them, whereas programs that provided minimal guidance left learners feeling aimless. Some participants stated that their program or school had done nothing in response to the pandemic, and that either their education had continued as usual or, in some instances, students were told to stay at home and self-direct their learning without any guidance from their program.

## Discussion

This study provides empirical data about the impact of COVID-19 on medical education around the world, notably the effects on learning and wellbeing. Taken together, our findings highlight how learners have been impacted regardless of their program’s response to the pandemic and whether or not they were removed from the clinical environment. However, the extent that COVID-19 has impacted learners differs significantly between levels of training and geographic regions.

COVID-19 has disrupted the traditional format and process of medical training for the vast majority of medical students and residents around the world. The rapid adoption of online learning by most schools may raise concerns about the quality of education (Mian & Khan 2020). Despite these concerns, equivalent educational outcomes between online and offline methods have been reported in the literature.(Pei & Wu 2019) The removal of learners from the clinical setting may diminish training (particularly for residents in time-based models of training), influencing clinical competence acquisition and career-decision making – all of which may affect patient care and health system outcomes (Goldhamer et al. 2020). While COVID-19 has required programs to adapt their curricula, this may be an opportunity to reconceptualize, innovate, and improve the current models of medical training, and ultimately catalyze broader educational transformation (Goldhamer et al. 2020; Sklar 2020).

Consistent with early reports, medical students feel under-utilized and have a strong desire to help out clinically during times of need (Lauren et al.; Herman et al. 2007; Gallagher & Schleyer 2020; Kalet et al. 2020; Li & Bailey 2020). However, not all learners may share these views. In addition to concerns about physical safety, some participants in our study believed the role of trainees is to be educated and that any deviation from that role is inappropriate. Others felt personally invested in contributing to care and viewed it as their personal calling and professional obligation. The optimal role of trainees during pandemics is complicated given their dual role as learners and healthcare providers – the extent to which each role is prioritized during emergencies remains debated. On one hand, it has been argued that medical students specifically, as an unpaid workforce, are not essential workers, and may increase disease transmission and waste scarce supplies of PPE (Menon et al. 2020). In contrast, it can also be argued that medical students are a semi-skilled workforce that can alleviate the burden on residents and staff, and should gain experience in preparation for future pandemics when they will no longer be trainees. In one study, hospital leaders viewed medical students who graduated early to help during COVID-19 as “game changers” (Flotte et al. 2020). It has also been argued that medical education is equally important to patient care during a pandemic (Klasen et al. 2020). This may be a unique opportunity to cultivate leadership skills if learners are well-supported (Brand 2020). For learners in the clinical environment, supervision, feedback, and evaluation as well as access to adequate wellness supports remain crucial (Brand 2020; Klasen et al. 2020). The ethical and practical considerations of utilizing medical learners during a pandemic remains an ongoing point of discussion and needs to be weighed against potential benefits of learning and implications to patient care (Simonds & Sokol 2008; Bauchner & Sharfstein 2020).

COVID-19 is expected to affect the mental health of the public(Venkatesh & Edirappuli 2020) as well as that of healthcare professionals (Lai et al. 2020). Previous health emergencies and disruptions to training, such as SARS and Hurricane Katrina, have been shown to diminish the mental health of trainees and healthcare professionals (Kisely et al. 2020). Our study provides similar evidence but from a global perspective that the wellbeing of medical learners has been negatively impacted by COVID-19 across all levels of training and geographic regions. This is concerning given the existing evidence on the alarming prevalence of burnout and psychiatric morbidities in learners, as COVID-19 may amplify these pre-existing issues (Puthran et al. 2016; Frajerman et al. 2019; Quek et al. 2019; Hartzband & Groopman 2020). Pandemic responses have superimposed new stressors while exacerbating existing stressors for learners. The “anticipatory loss” associated with the disruption of traditional processes and milestones (e.g., electives, residency match, licensing exams) appears associated with anxiety, and front-line learners (e.g., residents) may be particularly prone to wellness stressors (Gallagher & Schleyer 2020; Li & Bailey 2020). Suggestions for mitigating the psychological consequences of COVID-19 on front-line care providers include clear communication, education, and psychological support, among others (Kisely et al. 2020).

This pandemic remains an ongoing challenge for medical education. While the uncertainty of COVID-19 required organizations to adopt a “worst-case scenario” approach, medical schools not only continue to react to the needs of the health system, but proactively plan for the future. While some participants in this study applauded their program’s response, it is important to learn from this experience and improve planning and policies for current and future emergencies that may disrupt medical education (O’Byrne 2020). Recommendations are increasingly being discussed in the literature (The Coalition for Physician’s Accountability Work Group on Medical Students in the Class of 2021; Kachra & Brown 2020; Klasen et al. 2020; O’Byrne 2020; The Association of Faculties of Medicine of Canada 2020). Considerations for medical educators synthesized from our results and existing recommendations are summarized in Table 4. We must continue to share innovations and key lessons amongst the global medical education community. Future research is warranted to understand the long-term impact and potential consequences of the COVID-19 on education, patient care, and health system outcomes.

**Table 4.** Considerations for Medical Education During COVID-19

| Area | Considerations for Medical Schools and Residency Programs |
| --- | --- |
| Teaching and Learning | • Recognize that the rapid shift to online learning modalities likely requires ongoing evaluation, refinements, and improvement.<br>• Acknowledge that learners may have difficulties learning from home as well as challenges accessing a computer or the internet.<br>• Provide learners with self-directed learning resources with structure and guidance to promote student motivation and accountability.<br>• Simultaneously collect real-time feedback and use rapid-cycle improvement to promote continuous improvements while adapting new formats or processes.<br>• Identify potential innovations to maintain the fidelity and benefits of in-person learning, particularly for clinical skills.<br>• Consider integrating educational innovations created in response to COVID-19 into the core curriculum to be sustained after the pandemic.<br>• Scale what is working to other areas of your curriculum. Share success stories with others who may be struggling with similar challenges and could benefit from these strategies, particularly those in similar institutions (e.g., between residency programs).<br>• Conduct program evaluations of the new formats and processes adopted in response to COVID-19 to compare outcomes and provide evidence of merit and worth to learners and stakeholders. |
| Utilizing Learners | • Recognize that learners may be highly motivated to help out clinically during times of need and consider potential opportunities to engage them in meaningful roles – especially medical students when removed from their clinical rotations.<br>• Consider the training level, skillset, and experiences of learners. When appropriate, junior learners (e.g., medical students or junior residents) may be helpful to provide care in less acute settings or assist with low-risk roles to free senior residents and faculty to provide acute and intensive care.<br>• Provide learners working clinically with proper training on how to use personal protective equipment and provide care to COVID-19 patients.<br>• As learners have diverse needs, an individualized approach should be considered when engaging learners in the workforce – don’t assume all learners want to help out clinically. Learners may also be willing to help out in non-clinical ways, such as volunteering in the community.<br>• Continue providing learners with adequate supervision, feedback, and evaluation in the clinical setting as much as possible.<br>• Ensure safety equipment is available to learners in the clinical environments as well as health insurance if it is not already provided. |
| Supporting Learners | • Recognize the potential burden of COVID-19 on learner well-being and proactively provide systemic and programmatic support, appropriate access to counselling, peer support systems, and social networking.<br>• Acknowledge that the diverse and unique needs of learners may differ for some groups at this time, including students who may have a disability, parents, caregivers, and international learners<br>• Approach medical learner well-being holistically to ensure that mental, physical, intellectual, social, and occupational wellness is achieved.<br>• Cultivate a culture of wellness both systemically and programmatically (upstream) so that learners are able to thrive (downstream) during their training as well as disruptive events such as COVID-19.<br>• Recognize that “anticipatory loss” of milestones may be associated with learner anxiety and try to mitigate concerns relating to these areas, including: cancellation of electives, reduced clinical exposures, concerns about career decision making, anxiety about what specialties to apply to for residency with limited clinical exposure and the cancellation of electives, disruption of traditional timelines and processes, cancellation of licensing examinations, impact on career progression and obtaining employment, etc. |
| Communicating with Learners | • Communicate with learners clearly, early, and often – but not too often.<br>• Use centralized and streamlined modalities to reduce information overload, such as an e-mail update or frequently updated website, rather than social media. Consider surveying learners to ask about communication preferences and tailor the communication in response to their needs.<br>• Provide clear and concise updates to learners relating to the impacts of COVID-19 on their education, including changes to their program, timelines, and milestones.<br>• Be transparent about decision-making processes, outcomes, and contingency plans.<br>• Provide reassurance as much as possible and clearly communicate supports and resources to learners, such as institutional wellness resources, career counselling, academic advisors, peer supports, etc. |
| Policy, Planning, and Decision Making | • Engage learners in the decision-making process as much as possible, and as early as possible.<br>• Include learners on planning committees and in the development of policies.<br>• Create avenues for learners to be engaged in decision-making and to provide feedback to administrators and decision-makers, such as town halls, open office hours, or surveys.<br>• Acknowledge the diverse experiences and needs of learners and allow for flexibility in solutions. For instance, recognize that some – but not all – junior learners may wish to help and that the health system may benefit from utilizing this skilled workforce. Provide multiple options to learners when possible to promote autonomy and flexibility.<br>• Develop a robust policies and procedures from this experience. Plan for how the program or institution will respond to disruptions to medical training, including any subsequent waves of COVID-19, future pandemics, climate change emergencies, etc. |

### Limitations

We acknowledge that this study has several limitations, including the response and selection bias. The response rates from residents are low, potentially because they continued working clinically and the disruptions to their training were less extreme than for medical students who were completely removed from their training environments. Further, this study excluded participants who did not speak any of the 20 languages of the survey and there may be inconsistencies across the survey translations despite our best attempts to ensure questions and constructs translated accurately. Due to the public circulation of the survey, participants could not be validated as medical learners. A small fraction (3.5%) of participants did not specify their level of training and were excluded from the comparisons across levels of training. The geographic comparisons remain limited, as other classification systems (e.g., the World Health Organization regions) may have generated different results and there may be more pronounced differences between countries within similar regions (e.g., the United States vs. Canada). Finally, the variable responses to COVID-19 across countries and time is worth noting. Numerous qualitative comments highlighted how national responses informed the responses of professional organizations and training programs. Future research could examine potential associations between temporality and the political and health system responses to examine how these factors may extend to influence medical learners (Pablos-Méndez et al. 2020).

## Conclusions

This global study provides empirical data surrounding the profound impact that COVID-19 has had on medical learners, which can inform responses to the current pandemic, future disruptions to training, and future research. The results highlight the differences and similarities between levels of training and geographic regions. Despite the global shift toward online education, all education may not be created or perceived equally, as the results of this study suggest medical students fear their education will be of lower quality at this time. Both medical students and residents report their under-utilization, yet the role of the medical learner during a pandemic remains controversial. A delicate balance remains between medical training and the acute needs of the health system to ensure that both education and patient care remain safe. Finally, this study highlights how the COVID-19 pandemic has negatively impacted the wellbeing of medical learners globally, particularly those at the postgraduate level of training, which may be associated with their anxiety relating to the anticipatory losses of their educational experience. While the goal of this study was to broadly examine the impacts of COVID-19 on medical education, it highlights several areas for further exploration. Future research is warranted to better understand the implications of COVID-19 on learner outcomes and to generate evidence-based strategies surrounding how best to teach, engage, and support medical learners during similar events.

### Practice Points

- The rapid reorganization of medical training in response to COVID-19 has been substantial across all levels of training and geographic regions
- Medical learners are concerned that the disruption will negatively influence their learning, training trajectory, and career outcomes.
- The majority of medical learners feel under-utilized and are motivated to help out during times of need
- The mental health and wellbeing of learners may be further diminished at this time
- Medical schools should ideally learn from this experience and improve planning and policies for current and future disruptions or emergencies

## Data Availability

Data referred to in the manuscript is not publicly available at this time.

## Acknowledgments and Declaration of Interest

Notes on Contributors:

Allison Brown, PhD is an Assistant Professor in the Department of Medicine and Department of Community Health Sciences at the University of Calgary Cumming School of Medicine.

Aliya Kassam, PhD is an Assistant Professor in the Office of Postgraduate Medical Education and the Department of Community Health Sciences at the University of Calgary Cumming School of Medicine.

Mike Paget, BFA is a Manager of Academic Technologies in Undergraduate Medical Education at the University of Calgary Cumming School of Medicine.

Kenneth Blades, MA is a research analyst at the University of Calgary Cumming School of Medicine. Megan Mercia is a baccalaureate student in the Faculty of Science at the University of Calgary.

Rahim Kachra, MD EdM is an Associate Clinical Professor in the Department of Medicine and the Director of Teaching Innovations in Undergraduate Medical Education at the University of Calgary Cumming School of Medicine.

## Acknowledgements

The authors wish to acknowledge the O’Brien Institute for Public Health for funding this research, members of the University of Calgary medical education community for their support in conducting this study, as well as the Conjoint Health Research Ethics Board (Ashley Krecsy) for reviewing this study in a timely manner. Thanks to Kayla Atchison, Kyle Lafreniere, Suhair Bandeali, Krista Reich, Matthew Mancusco, Tefani Perera, and Nadiya Goswami for pilot testing the initial survey and providing feedback to help improve the survey items and functionality. We also wish to acknowledge Erika Tan and the Immigration Services Centre of Calgary and the assistance of numerous individuals who helped with the translation of the survey into additional languages: Philippe Pepin (French), Yusuf Yilmaz (Turkish), Dinesh Chavda (Hindi), Tayebah Chaudhry (Urdu), Giuliana and Salvatore Guarna (Italian), Michael Ji (Korean), Joska Eelsingh (Dutch), Laura Massignani (Spanish), Shenghao Huang (Chinese), Elzbieta Raniszewska (Polish), Florentino Guerra Filho (Portuguese), Tomoka Tsuhara and Lea Mezger (German), Hengameh Kheirkhah (Farsi), Atipong Pathanasethpong (Thai), Thi Hong An Do (Vietnamese), Vesta Seibel (Ukrainian), Alena Yakimenka (Russian), and Aito Ueno (Japanese). Next, our study team wishes to acknowledge all who amplified this study over social media or other forms of snowball sampling, including the Association for Medical Education in Europe (AMEE), AMOpportunities. Finally, our study team wishes to acknowledge the patient partners who reviewed the results of this research and the draft manuscript.

## Ethical Approval

This study was reviewed and approved by the University of Calgary Conjoint Research Ethics Board (File 20-0484) and follows the ethical conduct of the Tri-Council Policy Statement required by the Government of Canada and complies with the World Medical Association’s Declaration of Helsinki on Ethical Principles for Medical Research Involving Human Subjects. Participation in this study was voluntary and implied consent was obtained from all participants. Participants had the option to not answer any question or select “I prefer not to answer” if they wished to skip a question.

## References

Association of American Medical Colleges. 2020. Important Guidance for Medical Students on Clinical Rotations During the Coronavirus (COVID-19) Outbreak | AAMC [Internet]. [accessed 2020 Jul 14]. https://www.aamc.org/news-insights/press-releases/important-guidance-medical-students-clinical-rotations-during-coronavirus-covid-19-outbreak

Association of American Medical Colleges. 2020. Important Guidance for Medical Students on Clinical Rotations During the Coronavirus (COVID-19) Outbreak | AAMC [Internet]. [accessed 2020 Jul 14]. https://www.aamc.org/news-insights/press-releases/important-guidance-medicalstudents-clinical-rotations-during-coronavirus-covid-19-outbreak

Bai YM, Lin CC, Lin CY, Chen JY, Chue CM, Chou P. 2004. Survey of stress reactions among health care workers involved with the SARS outbreak. Psychiatr Serv. 55(9):1055–1057.

Bauchner H, Sharfstein J. 2020. A Bold Response to the COVID-19 Pandemic: Medical Students, National Service, and Public Health. JAMA - J Am Med Assoc. 323(18):1790–1791.

Brand PLP. 2020. COVID-19: a unique learning opportunity if the well-being of learners and frontline workers is adequately supported. Perspect Med Educ. 9(3):129–131.

Crawford BE, Kahn MJ, Gibson JW, Daniel AJ, Krane NK. 2008. Impact of Hurricane Katrina on medical student academic performance: The tulane experience. Am J Med Sci. 336(2):142–146.

DiCarlo RP, Hilton CW, Chauvin SW, Delcarpio JB, Lopez FA, McClugage SG, Letourneau JG, Smith R, Hollier LH. 2007. Survival and recovery: Maintaining the educational mission of the Louisiana State University School of Medicine in the aftermath of Hurricane Katrina. Acad Med. 82(8):745–756.

Flotte TR, Larkin AC, Fischer MA, Chimienti SN, DeMarco DM, Fan P-Y, Collins MF. 2020. Accelerated Graduation and the Deployment of New Physicians During the COVID-19 Pandemic. Acad Med. Publish Ah.

Frajerman A, Morvan Y, Krebs MO, Gorwood P, Chaumette B. 2019. Burnout in medical students before residency: A systematic review and meta-analysis. Eur Psychiatry. 55.

Gallagher TH, Schleyer AM. 2020. “We Signed Up for This!” - Student and Trainee Responses to the Covid-19 Pandemic. N Engl J Med [Internet]. 382(25):e96(1-3). nejm.org

Goldhamer MEJ, Pusic M V, Co JPT, Weinstein DF. 2020. Can Covid Catalyze and Educational Transformation? Competency-Based Advancement in a Crisis. N Engl J Med.:1–3.

Hartzband P, Groopman J. 2020. Physician Burnout, Interrupted. N Engl J Med [Internet].:1–2. nejm.org

Harvey A. 2020. Covid-19: medical students and FY1 doctors to be given early registration to help combat covid-19. BMJ [Internet]. 368(March):m1268. http://dx.doi.org/doi:10.1136/bmj.m1268

Herman B, Rosychuk RJ, Bailey T, Lake R, Yonge O, Marrie TJ. 2007. Medical students and pandemic influenza. Emerg Infect Dis. 13(11):1781–1783.

Kachra R, Brown A. 2020. The new normal: Medical education during and beyond the COVID-19 pandemic. Can Med Educ J.:4–6.

Kalet AL, Jotterand F, Muntz M, Thapa B, Campbell B. 2020. Hearing the Call of Duty: What We Must Do to Allow Medical Students to Respond to the COVID-19 Pandemic. Wis Med J.

Kassam A, Ellaway R. 2020. Acknowledging a Holistic Framework for Learner Wellness. Acad Med [Internet]. [accessed 2020 Jul 13] 95(1):9–10. http://journals.lww.com/00001888-202001000-00008

Kelley K, Clark B, Brown V, Sitzia J. 2003. Good practice in the conduct and reporting of survey research. Int J Qual Heal Care. 15(3):261–266.

Kisely S, Warren N, McMahon L, Dalais C, Henry I, Siskind D. 2020. Occurrence, prevention, and management of the psychological effects of emerging virus outbreaks on healthcare workers: rapid review and meta-analysis. BMJ. 369:m1642.

Klasen JM, Vithyapathy A, Zante B, Burm S. 2020. “The storm has arrived”: the impact of SARS-CoV-2 on medical students. Perspect Med Educ. 9(3):181–185.

Lai J, Ma S, Wang Y, Cai Z, Hu J, Wei N, Wu J, Du H, Chen T, Li R, et al. 2020. Factors Associated With Mental Health Outcomes Among Health Care Workers Exposed to Coronavirus Disease 2019. JAMA Netw open. 3(3):e203976.

Landis MS, Bradley JW. 2005. The Impact of the 2003 SARS Outbreak on Medical Students at the University of Toronto. Univ Toronto Med … [Internet].:0–6. https://pdfs.semanticscholar.org/c574/8067e223ea25670788e0a4ca13d0771dccd3.pdf

Lauren G, Paige K, Pardis P, Terry W, D GJ. Author: Long Nathaniel; Wolpaw Daniel R. MD; Boothe David; Caldwell Catherine; Dillon Peter MD; Gottshall Lauren; Koetter Paige; Pooshpas Pardis; Wolpaw Terry MD, MHPE; Gonzalo Jed D. MD, MSc.

Li HO-Y, Bailey AMJ. 2020. Medical Education Amid the COVID-19 Pandemic. Acad Med. Published.

Marteau TM, Bekker H. 1992. The development of a six‐item short‐form of the state scale of the Spielberger State—Trait Anxiety Inventory (STAI). Br J Clin Psychol [Internet]. [accessed 2020 Jul 13] 31(3):301–306. https://onlinelibrary.wiley.com/doi/abs/10.1111/j.2044-8260.1992.tb00997.x

Maunder R, Hunter J, Vincent L, Bennett J, Peladeau N, Leszcz M, Sadavoy J, Verhaeghe LM, Steinberg R, Mazzulli T. 2003. The immediate psychological and occupational impact of the 2003 SARS outbreak in a teaching hospital. Can Med Assoc J. 168(10):1245–1251.

Menon A, Klein EJ, Kollars K, Kleinhenz ALW. 2020. Medical Students Are Not Essential Workers: Examining Institutional Responsibility During the COVID-19 Pandemic. Acad Med.

Mian A, Khan S. 2020. Medical education during pandemics: A UK perspective. BMC Med. 18(1):18–19.

Miles MB, Huberman AM, Saldaña J.. 2014. Qualitative data analysis: a methods sourcebook. [place unknown].

O’Byrne L. 2020. Medical students and COVID-19: the need for pandemic preparedness. J Med Ethics.:medethics-2020-106353.

Pablos-Méndez A, Vega J, Aranguren FP, Tabish H, Raviglione MC. 2020. Covid-19 in Latin America. BMJ [Internet]. [accessed 2020 Jul 27] 370. https://www.bmj.com/content/370/bmj.m2939

Pei L, Wu H. 2019. Does online learning work better than offline learning in undergraduate medical education? A systematic review and meta-analysis. Med Educ Online.

Puthran R, Zhang MWB, Tam WW, Ho RC. 2016. Prevalence of depression amongst medical students: A meta-analysis. Med Educ. 50(4).

Quek TTC, Tam WWS, Tran BX, Zhang M, Zhang Z, Ho CSH, Ho RCM. 2019. The global prevalence of anxiety among medical students: A meta-analysis. Int J Environ Res Public Health. 16(15).

Rambaldini G, Wilson K, Rath D, Lin Y, Gold WL, Kapral MK, Straus SE. 2005. The impact of severe acute respiratory syndrome on medical house staff: a qualitative study. J Gen Intern Med [Internet]. [accessed 2020 Jul 25] 20(5):381–5. http://www.ncbi.nlm.nih.gov/pubmed/15963157

Rieder MJ, Salvadori M, Bannister S, Kenyon C. 2004. Collateral damage: the effect of SARS on medical education. Clin Teach. 1(2):85–89.

Rose S. 2020. Medical Student Education in the Time of COVID-19. J Am Med Assoc. 323(21):2131–2132.

Sherbino J, Atzema C. 2004. “SARS-Ed”: Severe acute respiratory syndrome and the impact on medical education. Ann Emerg Med. 44(3):229–231.

Simonds AK, Sokol DK. 2008. Lives on the line? Ethics and practicalities of duty of care in pandemics and disasters. Eur Respir J. 34(2):303–309.

Sklar DP. 2020. COVID-19: Lessons From the Disaster That Can Improve Health Professions Education. Acad Med [Internet]. http://www.ncbi.nlm.nih.gov/pubmed/32544103

The Association of Faculties of Medicine of Canada. 2020. Ten Guiding Principles for Medical Education [Internet]. [accessed 2020 Jul 14]. https://afmc.ca/sites/default/files/pdf/2020-Ten_Guiding_Principles_for_Medical_Education_COVID_Era_en.pdf

The Coalition for Physician’s Accountability Work Group on Medical Students in the Class of 2021. Final Report and Recommendations for Medical Education Institutions of LCME-Accredited, U.S. Osteopathic, and Non-U.S. Medical School Applicants [Internet]. [place unknown]; [accessed 2020 Jul 14]. https://www.aamc.org/system/files/2020-05/covid19_Final_Recommendations_ExecutiveSummary_Final_05112020.pdf

Venkatesh A, Edirappuli S. 2020. Social distancing in covid-19: What are the mental health implications? Br Med [Internet]. 369(April):2020. http://dx.doi.org/doi:10.1136/bmj.m1379

World Bank. How does the World Bank classify countries? [Internet]. [accessed 2020 Jul 13]. https://datahelpdesk.worldbank.org/knowledgebase/articles/378834-how-does-the-world-bankclassify-countries

